# Genetic Variants and Therapeutic Approaches in Maturity-Onset Diabetes of the Young: A Retrospective Analysis

**DOI:** 10.1101/2024.05.20.24307619

**Authors:** Lily Deng, Amy S. Shah, Mansa Krishnamurthy

**Affiliations:** Division Endocrinology, Department of Pediatrics, Cincinnati Children’s Hospital Medical Center, Cincinnati, OH, University of Cincinnati College of Medicine, Cincinnati, OH

**Keywords:** MODY, GLP-1, Genetics, Sulfonylurea

## Abstract

**Context:** Identifying Maturity-Onset Diabetes of the Young (MODY) in patients with diabetes is essential because treatment differs significantly from other forms of diabetes. We identified patients with MODY gene variants and evaluated their clinical characteristics and responses to treatment.

**Evidence Acquisition:** We identified 106 patients with genetic MODY variants. Demographics, islet autoantibodies at diabetes diagnosis, co-morbidities, and response to treatment by genetic variant were evaluated.

**Evidence Synthesis:** Patients diagnosed with MODY variants comprised 4% of the total population with diabetes. Mean age and HbA1c of patients with MODY at diagnosis were 10.5 years and 8.2%, respectively. Surprisingly, diabetic ketoacidosis was a presenting feature for some (n=7, 6.8%), and others with MODY had positive islet cell autoantibodies (n= 7, 6.6%). Variants in HNF1A, GCK, and HNF1B were frequently observed (20%, 22%, and 17% respectively), while rare variants in PDX1, RFX6, BLK, and CNOT1 were uncovered. Initial and follow up treatment of patients with MODY were compared. For each medication (Insulin, Metformin, Sulfonylureas, and GLP-1 receptor agonists), a reduction in HbA1c was observed at follow-up (0.3-21%). Insulin and sulfonylureas were associated with an increase in average BMI (insulin: +8.23%, n=21, sulfonylurea: +0.63%, n=12) at follow-up, metformin was intermediate (−2.46%, n=4), and GLP-1 receptor agonists demonstrated the greatest decrease in BMI (−4.79%, n=4).

**Conclusions:** The presence of islet autoantibodies or diabetic ketoacidosis does not preclude the diagnosis of MODY. Given the observed improvements in BMI and HbA1c, further investigation into the use of GLP-1 receptor agonists as treatment for MODY should be considered.

## Introduction

Maturity-onset diabetes of the young (MODY) is a monogenic form of diabetes mellitus that is dominantly inherited and accounts for approximately 1-5% of the pediatric population with diabetes^1–3^. Patients often present before 25 years of age with lack of beta cell autoimmunity and some residual beta cell function^4–6^. To date, 14 causative MODY genes have been identified^7^ with the most common being glucokinase (GCK), hepatocyte nuclear factor-1 alpha (HNF1A), and hepatocyte nuclear factor 4 alpha (HNF4A)^2,8^. Variants in these genes can lead to defects^6^ in glucose sensing and insulin secretion^3^. Clinical phenotypes differ based on the genetic defect, and treatment and management options for patients considerably differ from those with a diagnosis of type 1 or type 2 diabetes. For example, patients diagnosed with GCK pathogenic variants do not require medical management^8^, while patients with HNF1A and HNF1 homeobox B (HNF1B) pathogenic variants are usually responsive to sulfonylureas^9,10^. The recognition of MODY is crucial; unfortunately, MODY is commonly misdiagnosed, leading to unnecessary insulin treatment for numerous patients. Moreover, there is often a delay in correct genetic diagnosis from the initial diabetes diagnosis due to lack of clinical recognition or access to genetic testing.

Patients with MODY may have additional clinical findings due to their underlying genetic variants which further raises the importance of timely, accurate identification of this patient population. Renal cysts are found in patients with HNF1B pathogenic variants, exocrine pancreatic insufficiency in patients with carboxyl ester lipase (CEL) pathogenic variants, and optic nerve atrophy, hearing loss and neurodegeneration in patients with wolframin ER transmembrane glycoprotein (WFS1) pathogenic variants. Thus, timely genetic diagnosis tailors the most appropriate treatment of diabetes and allows for effective management of co-morbidities in this patient population^11^. Timely diagnosis of MODY also results in decreased healthcare costs and increased quality of life (due to medical management for MODY or cessation of unneeded medications) in a simulated model^12^.

Traditionally, treatment modalities for MODY have included lifestyle modifications, sulfonylureas, and insulin therapy. However, glucagon-like peptide-1 (GLP-1) receptor agonists have emerged as potential therapeutic options for the treatment of MODY^4,13–15^. Randomized, double-blinded clinical trials and case reports have shown a reduction in fasting plasma glucose levels with less frequent episodes of hypoglycemia^14^ and reduced hemoglobin A1c (HbA1c)^16^ in patients with HNF1A variants, while a significant reduction in HbgA1c was observed in a father-son cohort with a variant in HNF4A^13^ with another case report showing reduction in HbA1c and body mass index (BMI) in an 18-year-old with HNF1B-MODY^17^. GLP-1 receptor agonists exert an insulinotropic effect in a strictly glucose-dependent manner resulting in a low risk for hypoglycemia which is commonly seen as a side effect in insulin and sulfonylurea therapy^14^. Moreover, GLP-1 receptor agonists have been demonstrated to decrease BMI in pediatric and adult patients with type 2 diabetes^18,19^, suggesting that improvements in glucose tolerance and insulin sensitivity may have added advantages when using GLP-1 receptor agonists in the population with MODY.

The aim of our study was to characterize our patient population with MODY based on genetics and evaluate presentation, presence of co-morbidities and islet cell autoantibodies, and treatments at diagnosis and over time.

## Research Design and Methods

A retrospective chart review, utilizing diagnoses codes for MODY and MODY panel testing, was performed on patients diagnosed with diabetes at Cincinnati Children’s Hospital Medical Center (CCHMC) from January 2010 until June 2023. This revealed 106 patients with variants in known MODY genes. Thirty-six (36) of these patients had variants of unknown significance (VUS), 2 patients had possible pathogenic variants, and 19 patients had likely pathogenic variants. Thirty-nine (39) patients had pathogenic variants, and 1 patient had a variant originally designated as pathogenic but now classified as a polymorphism. The remaining genetic variants did not have documented clinical significance (1 patient was diagnosed with HNF1A pathogenic variant based on biopsy findings and staining.) Patients with only clinical findings suggestive MODY were not included in this study.

Genetic testing was performed in a clinical setting and included the following tests: Invitae monogenic diabetes panel, Prevention genetics, Seattle Children’s Hospital, and Gene Dx monogenic diabetes panels. Invitae utilizes next-generation sequencing technology (NGS) for both sequencing analysis and deletion/duplication analysis to analyze regions of each gene tested. This panel includes ATP binding cassette subfamily C member 8 (ABCC8), adaptor protein, phosphotyrosine interacting with PH domain and leucine zipper 1 (APPL1), B Lymphocyte Kinase (BLK), eukaryotic translation initiation factor 2 alpha kinase 3 (EIF2AK3), forkhead box P3 (FOXP3), GATA binding protein 4 (GATA4), GATA binding protein 6 (GATA6), GCK, GLIS family zinc finger 3 (GLIS3), HNF1A, HNF1B, HNF4A, immediate early response 3 interacting protein 1 (IER3IP1), insulin (INS), potassium inwardly rectifying channel subfamily J member 11 (KCNJ11), KLF transcription factor 11 (KLF11), motor neuron and pancreas homeobox 1 (MNX1), neuronal differentiation 1 (NEUROD1), neurogenin 3 (NEUROG3), NK2 homeobox 2 (NKX2-2), paired box 4 (PAX4), pancreatic and duodenal homeobox 1 (PDX1), peroxisome proliferator activated receptor gamma (PPARG), pancreas associated transcription factor 1a (PTF1A), regulatory factor x6 (RFX6), solute carrier family 19 member 2 (SLC19A2), WFS1, and ZFP57 zinc finger protein (ZFP57). The lab then uses Moon software tool to analyze exomes, supported by a gene-disease database, Apollo, to evaluate variants and determine clinical significance. Sherloc, Ivitae’s own variant classification algorithm includes data from their functional modeling platform and RNA analysis. If variants are re-classified, addendums are added to prior reports. The Prevention Genetics MODY panel utilizes NGS to target coding regions of each gene in addition to margins of 10 bases of noncoding DNA on either side of the targeted coding regions. Copy number variations (CNVs) can be detected with their technology, although if it is not technically possible to confirm smaller CNVs, then those are not included on the result report. Variants are described using Human Gene Variation Society recommendations. Genes tested include ABCC8, APPL1, BLK, CEL, CCR4-NOT transcription complex subunit 1 (CNOT1), GCK, glutamate dehydrogenase 1 (GLUD1), hydroxyacyl-CoA dehydrogenase (HADH), HNF1A, HNF1B, HNF4A, INS, KCNJ11, KLF11, NEUROD1, PAX4, PDX1, RFX6, and WFS1. Seattle Children’s MODY panel utilizes NGS technology to sequence target regions and a margin of at least 10 base pairs of introns surrounding the target region. Their methods can detect CNVs and small deletions or insertions. Genes included in this panel include ABCC8, APPL1, BLK, CEL, GCK, HNF1A, HNF1B, HNF4A, INS, INS-IGF2 readthrough (INS-IGF2), KCNJ11, KLF11, NEUROD1, PAX4, and PDX1. GeneDx utilizes NGS with CNV calling to evaluate for sequence variants in ABCC8, APPL1, BLK, CEL, GCK, GLUD1, HADH, HNF1A, HNF1B, HNF4A, INS, KCNJ11, KLF11, NEUROD1, PAX4, PDX1, RFX6, and WFS1. They mainly report pathogenic variants, likely pathogenic variants, and VUS. Genetic variants are classified according to clinical information provided by the ordering provider, Human Gene Mutation Database/other databases, phenotype, severity of sequence change and function, and population frequency.

Data reviewed from medical charts included patient demographics (age, race, sex/ethnicity), anthropometrics (including BMI), laboratory data, genetic testing results, initial diabetes presentation, and medication use.

## Statistics and Data Analysis

Statistical analyses were performed using GraphPad Prism 9.0. If the HbA1c value was reported as >14%, a value of 14% was used in the analysis as there was no exact HbA1c value available due to the lab assay performed. For patients receiving a medication and then concurrently starting a second medication, their BMI and HbA1c parameters were assessed at initiation and follow-up of the first medication. Follow-up was defined as the first visit documented since initiation of medication, which ranged from 1-9 months with majority of follow ups between 1-3 months. This was counted separately from their BMI and HbA1c at the initiation and follow-up of the second medication. Changes in BMI and HbA1c were measured by calculating percent difference between initial BMI or HbA1c and follow-up BMI or HbA1c, respectively. Several patients on medications did not have follow ups within the specified time for follow up (above), so their BMI and HbA1c data were not included to limit confounding. One-way analysis of variance (ANOVA) followed by Tukey’s post hoc test was used to evaluate the differences in BMI and HbA1c between medication classes at initial diabetes diagnosis and follow-up. A p value of <0.05 was considered significant.

## Results

### Baseline Patient Characteristics

We identified 106 patients with variants in known MODY genes (Figure 1, Supplement Table 1). Nine of these patients had benign genetic variants or variants not associated in MODY genes. Forty of these patients had variants of unknown significance, two patients had possibly pathogenic variants, and nineteen patients had likely pathogenic variants. Thirty-five patients had pathogenic variants. Patients with any variant in a MODY gene comprised 4.0% of the total patient population with diabetes at our center receiving care during the same interval (Supplement Figure 1). Pathogenic variants and VUS in GCK, HNF1A, HNF4A, and HNF1B comprised a total of around 66% of those found in our population with MODY. Moreover, the majority of pathogenic (83%) and likely pathogenic variants (77%) (Supplement Table 2) were found within the cohort of patients with GCK, HNF1A, HNF4A, and HNF1B variants. Rarer pathogenic variants and VUS were also identified in APPL1, BLK, PDX1, CNOT1, NEUROD1, and RFX6 (Supplement Table 1).

**Figure 1.**
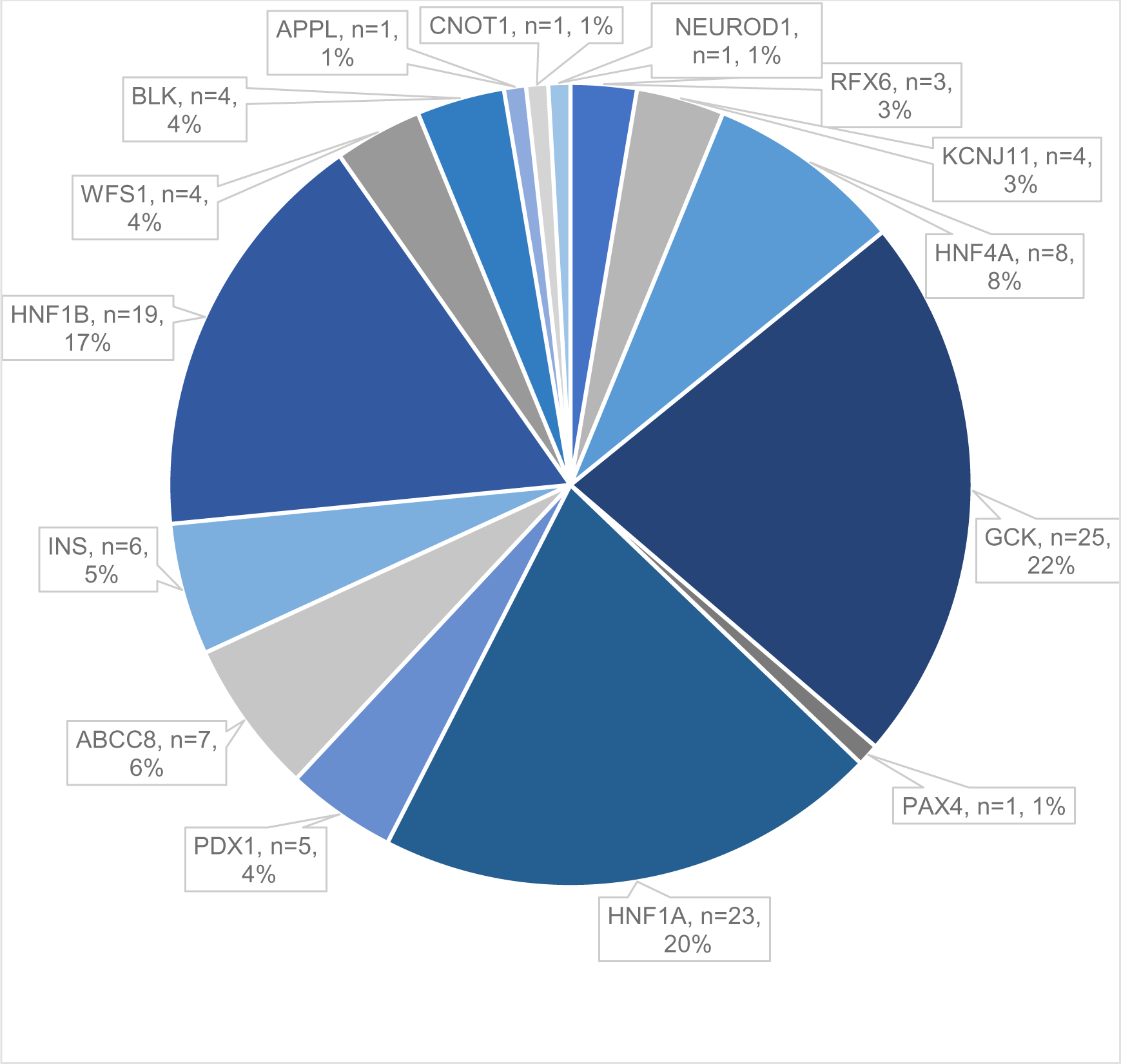
Pathogenic variants and variants of unknown significance categorized based on gene. Number of patients listed along with percentage within our MODY population.

The average mean age ± standard deviation (SD) at diabetes diagnosis was 10.5 ± 5.7 years, and the mean HbA1c at diagnosis was 8.2% ± 3.2 for our population with MODY. Sex distribution of our population with MODY was 55.7% female and 44.3% male (Table 1). Most patients with MODY self-reported race/ethnicity as non-Hispanic white (66%). MODY was also diagnosed in patients who self-identified as Hispanic, non-Hispanic Black, and Asian/Pacific Islander. Negative islet cell autoantibodies were observed in 62.3% of patients with MODY, but 6.6% of patients had at least one islet cell autoantibody prior to the initiation of insulin therapy (including anti-glutamic acid decarboxylase (GAD) and zinc transporter 8 (ZnT8)). One patient was positive for two islet autoantibodies (with a likely pathogenic GCK variant), while another patient (with a variant in GCK) tested positive for ZnT8. Moreover, 31.1% of the total patient population with MODY did not have islet cell autoantibody screening collected at time of diabetes diagnosis (perhaps as they were suspected to have MODY from diabetes onset).

**Table 1.** Clinical Characteristics of all MODY patients at Diabetes Diagnosis and Treatment at Diagnosis and Co-morbidities of all MODY patients at Diabetes Diagnosis

| Characteristics of All Patients at Diabetes Diagnosis and Treatment at Diagnosis |  |  |  |  |
| --- | --- | --- | --- | --- |
|  | n | Percent | Mean | SD |
| **Age of Diabetes Diagnosis (years)** | 106 |  | 10.51 | 5.69 |
| **BMI At Diabetes Diagnosis (m/kg^2)** | 106 |  | 22.39 | 6.13 |
| **HbA1c At Diabetes Diagnosis (%)** | 106 |  | 8.20 | 3.17 |
| **Sex** | 106 |  |  |  |
| Female | 59 | 55.66% |  |  |
| Male | 47 | 44.34% |  |  |
| **Race** | 106 |  |  |  |
| White (non-Hispanic) | 70 | 66.04% |  |  |
| White (Hispanic) | 11 | 10.38% |  |  |
| Black | 19 | 17.92% |  |  |
| Asian/Pacific Islander | 3 | 2.83% |  |  |
| Unknown | 3 | 2.83% |  |  |
| **Islet Cell Autoantibody Screen** | 106 |  |  |  |
| Positive | 7 | 6.60% |  |  |
| Negative | 66 | 62.26% |  |  |
| Not obtained | 33 | 31.13% |  |  |
| **Initial Diagnosis** | 99 |  |  |  |
| Type 1 DM | 36 | 36.36% |  |  |
| Type 2 DM | 7 | 7.07% |  |  |
| Beta cell mismatch hypoglycemia | 1 | 1.01% |  |  |
| Neonatal diabetes | 1 | 1.01% |  |  |
| MODY | 28 | 28.28% |  |  |
| Agenesis of the pancreas | 2 | 2.02% |  |  |
| Combination | 24 | 24.24% |  |  |
| **Initial Therapy** | 100 |  |  |  |
| Insulin Alone | 36 | 36.00% |  |  |
| MDI | 35 |  |  |  |
| 70/30 Insulin | 1 |  |  |  |
| Metformin | 7 | 7.00% |  |  |
| Sulfonylurea | 1 | 1.00% |  |  |
| Insulin and Metformin | 3 | 3.00% |  |  |
| None | 53 | 53.00% |  |  |
| **Co-morbidities of All Patients at Diabetes Diagnosis** |  |  |  |  |
|  | **N** | **Percent** |  |  |
| **Blood Pressure** | 79 |  |  |  |
| Normal | 73 | 92.41% |  |  |
| Hypertensive (mild to Stage I) | 6 | 7.59% |  |  |
| **Acanthosis** | 103 |  |  |  |
| Present | 15 | 14.56% |  |  |
| Absent | 88 | 85.44% |  |  |
| **Ketoacidosis At Diagnosis** | 103 |  |  |  |
| Present | 7 | 6.80% |  |  |
| Absent | 96 | 93.20% |  |  |
| **Lipid Profile** | 106 |  |  |  |
| Normal | 47 | 44.34% |  |  |
| Abnormal | 24 | 22.64% |  |  |
| Not obtained | 35 | 33.02% |  |  |
| **TSH** | 106 |  |  |  |
| Normal | 67 | 63.21% |  |  |
| Abnormal | 6 | 5.66% |  |  |
| Not obtained | 33 | 31.13% |  |  |
| **Celiac Screening** | 100 |  |  |  |
| Negative | 43 | 43.00% |  |  |
| Positive | 0 | 0.00% |  |  |
| Not obtained | 57 | 57.00% |  |  |

At diagnosis of diabetes, the most common initial diagnoses assigned to these patients were type 1 diabetes (36%), MODY (28%), unknown diabetes type (24%), and Type 2 diabetes (7%) (Table 1). The most common initial therapy at diagnosis of diabetes was no medication, followed by basal bolus insulin therapy (Table 1).

### Genetic Characteristics of Patient Population

By gene, patients with a variant in HNF1B had the shortest average duration from diagnosis of diabetes to diagnosis of MODY, -7.2 months (Table 2) as n=10/20 of these patients were diagnosed with MODY prior to developing diabetes when renal cysts or other malformations were identified *in utero* or in early childhood. There was no significant difference across genetic variants when duration to MODY diagnosis was analyzed (p=0.1.) The average age at diagnosis of diabetes by gene varied from 3 to 16 years (Table 2). HbA1c at diabetes diagnosis was highest in a patient with an APPL1 (n=1) variant, followed by a NEUROD1 (n= 1) variant and a patient with an INS (n=1/6) variant.

**Table 2:**
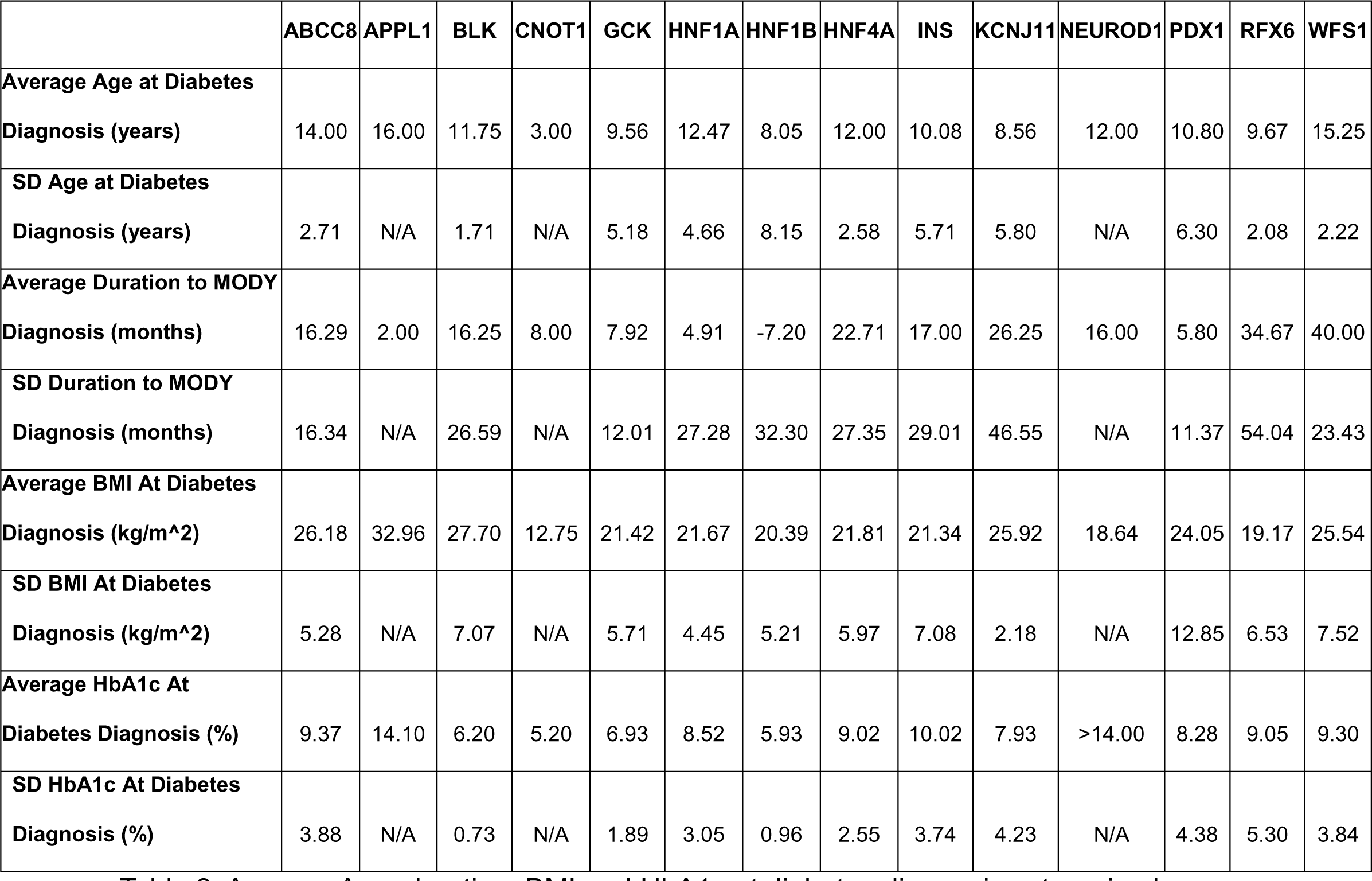
Average Age, duration, BMI and HbA1c at diabetes diagnosis categorized based on gene.

### Comorbidities in Patient Population

Comorbidities were also evaluated for each patient at time of diabetes diagnosis. Most patients (92.4%) had a normal systolic and diastolic blood pressure, and 7.6% of patients were diagnosed with elevated blood pressure or stage 1 hypertension (Table 1). Acanthosis nigricans was noted on physical exam in 14.6% of patients. Diabetic ketoacidosis was a presenting feature in 6.8% of patients. These patients were found to have VUS in GCK, KCNJ11, RFX6, INS, ABCC8, WFS1, and HNF1A. Moreover, lipid profile, thyroid stimulating hormone (TSH), and celiac screening were obtained at diabetes diagnosis (as patients were presumed to have a diagnosis of type 1 diabetes) in 67%, 69%, and 43% of the patient population with MODY, respectively (Table 1). Abnormal lipids (elevated triglycerides, low HDL, elevated LDL, or elevated total cholesterol) were found in 22.6% of patients, 5.7% had slightly abnormal TSH, and no patients screened positive for celiac disease.

### Initial and Current Treatment Modalities Utilized in Patients with MODY

The initial and current treatment modalities in our patient population with MODY were evaluated along with change in BMI and HbA1c from diagnosis to follow-up. No medications at diagnosis were administered in 53% of MODY patients, and 53% were not on medications at follow-up (Tables 1 and 3). Of the 47% of the patients who received medications at diagnosis, treatments included insulin (36%), metformin (7%), insulin and metformin combination (3%), and sulfonylureas (1%) (Table 1.) The therapies during the 1-9 months of follow-up included insulin (21.6%), sulfonylureas (15.9%), GLP-1 receptor agonists (4.6%), and metformin (4.6%) (Table 3.) The follow-up visit utilized for this data was the first visit the patient returned after starting the medication.

**Table 3:** Current treatment of MODY patients categorized by genetic variant. *Patients with more than 1 genetic variant. The patient with a variant in PAX4 was on acarbose (which is not listed in this table category.)

| Genetic Variant | Sulfonylurea | GLP-1 receptor agonist | Insulin alone | Metformin | Combination | No Medication |
| --- | --- | --- | --- | --- | --- | --- |
| **ABCC8** | 3 (37.5%) | 0 (0%) | 0 (0%) | 3 (37.5%) | 2* (25%) | 0 (0%) |
| **APPL1** | 0 (0%) | 0 (0%) | 0 (0%) | 0 (0%) | 1 (100%) | 0 (0%) |
| **BLK** | 0 (0%) | 1* (25%) | 1* (25%) | 1 (25%) | 0 (0%) | 1 (25%) |
| **CNOT1** | 0 (0%) | 0 (0%) | 0 (0%) | 0 (0%) | 0 (0%) | 1 (100%) |
| **GCK** | 1 (3.85%) | 0 (0%) | 2 (7.69%) | 2 (7.69%) | 0 (0%) | 21 (80.77%) |
| **HNF1A** | 8 (36.36%) | 0 (0%) | 6 (27.27%) | 0 (0%) | 2 (9.09%) | 6* (27.27%) |
| **HNF1B** | 0 (0%) | 1 (5%) | 1 (5%) | 0 (0%) | 0 (0%) | 18 (90%) |
| **HNF4A** | 2* (22.22%)1 | 0 (0%) | 4 (44.44%) | 0 (0%) | 2* (22.22%) | 1* (11.11%) |
| **INS** | 0 (0%) | 1 (16.67%) | 4* (66.67%) | 0 (0%) | 1* (16.67%) | 0 (0%) |
| **KCNJ11** | 1 (25%) | 1* (25%) | 1* (25%) | 0 (0%) | 0 (0%) | 1 (25%) |
| **NEUROD1** | 0 (0%) | 0 (0%) | 1 (100%) | 0 (0%) | 0 (0%) | 0 (0%) |
| **PAX4\*\*** | 0 (0%) | 0 (0%) | 0 (0%) | 0 (0%) | 0 (0%) | 0 (0%) |
| **PDX1** | 0 (0%) | 0 (0%) | 2 (50%) | 0 (0%) | 0 (0%) | 2* (50%) |
| **RFX6** | 0 (0%) | 0 (0%) | 2* (66.67%) | 0 (0%) | 0 (0%) | 1 (33.33%) |
| **WFS1** | 0 (0%) | 0 (0%) | 2 (50%) | 1 (25%) | 0 (0%) | 1 (25%) |

Patients with variants in HNF1A comprised the largest group treated with sulfonylureas at follow-up (36% of the total population with HNF1A variants) (Table 3). Patients with HNF1A, HNF4A, and INS variants comprised the largest group treated with insulin therapy (27%, 44%, and 67% of the population with those variants respectively) (Table 3). Regardless of medication class, the mean change in HbA1c at follow-up was - 1.2%, with the highest reduction seen in patients on insulin (−2.7%.) Patients on insulin (n=21) also experienced a mean increase in weight of 8.2% whereas patients on other medications including metformin and GLP-1 receptor agonists (Semaglutide and Liraglutide) were noted to have weight loss (−2.5% and -4.8% respectively) (Figure 2). Compared to patients on GLP-1 receptor agonists, patients on insulin alone had a significant increase (p=0.24) in BMI at follow-up (Figure 2). Patients on sulfonylureas (n=12) had weight gain at follow up (0.63%) (Figure 2). The greatest percent in BMI reduction was seen in patients on GLP-1 receptor agonists (−4.8%, n=4) (Figure 2).

**Figure 2.**
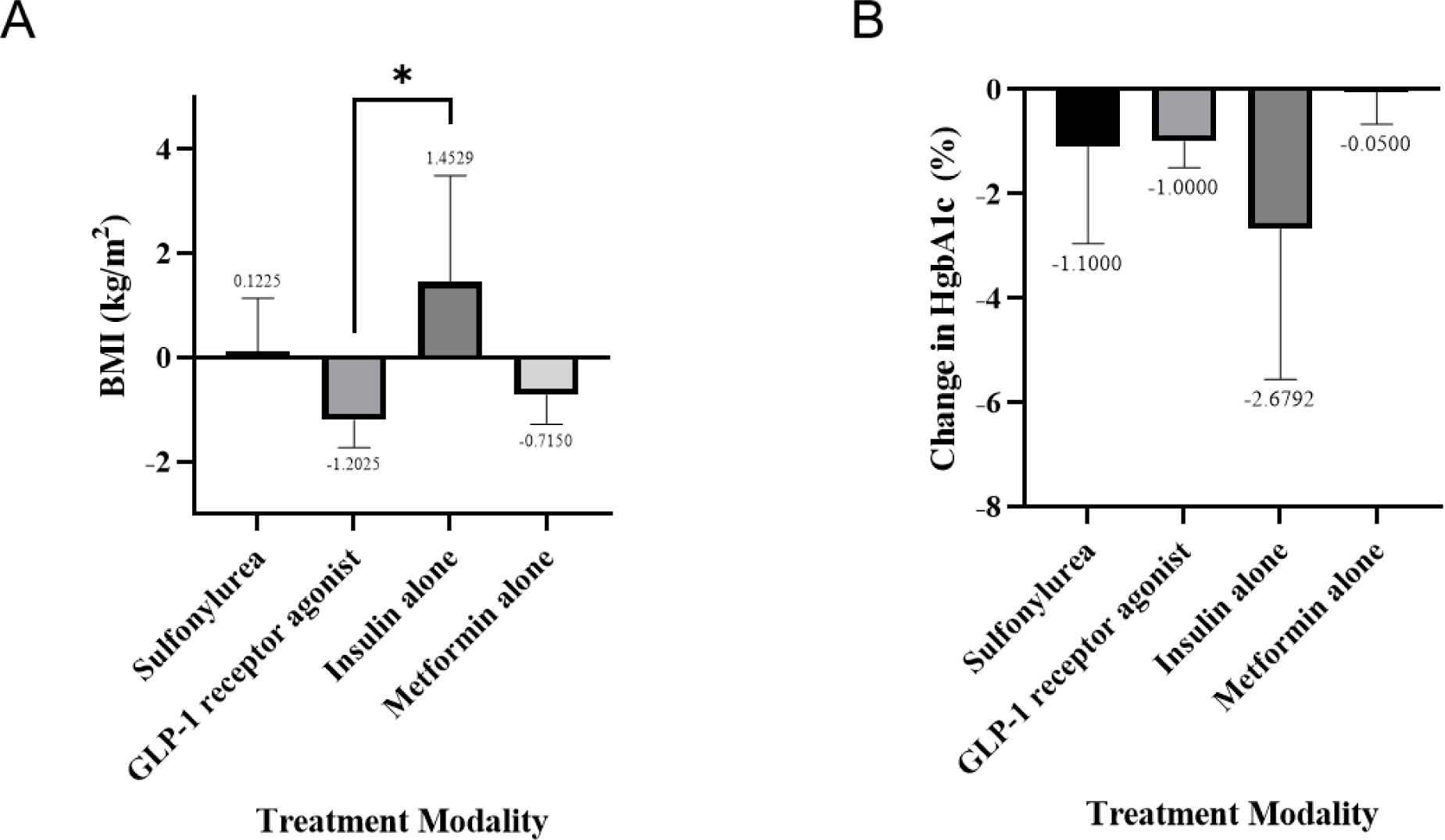
A,B. Change in BMI and HbA1c categorized by medication. *p value < 0.05. The following data was not included in assessment of medication response: patients with follow-up greater than 9 months after initiation of medication, patients without records of initial presentation data or medication use, patients without follow-up data and patients concurrently receiving two or more medications.

## Discussion

In this retrospective study, we performed an in-depth characterization of our patient population with genetically confirmed MODY to evaluate demographics, laboratory data, co-morbidities and treatment modality at diagnosis and current treatments. Our study provides clear support for identifying patients with MODY: most patients are non-Hispanic white, test negative for islet cell antibodies, have mild hyperglycemia reflected by a median HbA1c at presentation of 6.7%, and have a BMI <85^th^ percentile for age and sex. Pathogenic genetic variants were more commonly found in GCK, HNF1A, and HNF1B; several rare variants were also uncovered in genes including APPL, BLK, PDX1, RFX6, CNOT1, PAX4, and NEUROD1. Finally, a reduction in BMI was noted in all treatment groups except for insulin, while the greatest reduction in BMI was noted in the group receiving GLP-1 receptor agonist therapy. Most of the patients on sulfonylureas had mild elevations in BMI at follow-up, except one patient with excellent BMI reduction due to lifestyle modifications.

While several of the findings of our study are consistent with the literature, there are several differences to highlight. MODY has traditionally been described to occur more commonly in non-Hispanic white youth^20,21^. While our data is consistent with this, we also noted MODY in several other race/ethnic groups including Hispanic, non-Hispanic Black, and Asian/Pacific Islander youth. Moreover, the percentage of patients with positive islet cell autoantibodies in our population with MODY (6.6%) is higher than previously reported in patients with MODY (<1%)^22^. The co-existence of positive islet autoantibodies and genetic variants consistent with MODY diagnosis have previously been reported by other groups^23,24^. A study in 28 Czech patients demonstrated that 25% of their patients with MODY had positive islet autoantibodies ^24^. Taken together with our findings, the presence of islet cell antibodies does not completely preclude the diagnosis of MODY. Whether the presence of islet cell antibody influences the rate of beta cell decline due to future autoimmunity are areas for future work.

We would also like to note that a percentage of the patients (6.8%) with MODY, with negative islet cell antibodies, presented in diabetic ketoacidosis at diabetes diagnosis. Interestingly, one of these patients had a VUS in GCK, and it has been widely reported in the literature that patients with GCK-MODY are mostly asymptomatic with mild non-progressive hyperglycemia ^25^. No additional genetic variants in known MODY genes were found in this patient, and islet cell antibodies were also negative. This contrasts with most of the literature on MODY, that patients present with lower HbA1c and that diabetic ketoacidosis is exceedingly rare. Our data indicate that the presentation of diabetic ketoacidosis does not preclude the diagnosis of MODY and that thorough evaluation for MODY should still be performed, if suspected, regardless of initial presentation of diabetes.

Interestingly, not all of our patients diagnosed with MODY were screened at diagnosis of diabetes for co-morbidities. This could be as physicians suspected MODY at diagnosis whereas others who suspected type 1 diabetes obtained the standard screenings per our institutional protocol. This could also be due to the large number of patients with GCK variants, which have been noted to rarely develop microvascular complications^26^. Furthermore, given that the diagnosis of MODY is due to genetic variants and not autoimmunity, there is less association with other autoimmune diseases such as thyroid disease and celiac disease, which are typically screened in patients with autoimmune type 1 diabetes. Consequently, these screenings were not obtained in some of our patient population. Thus, genetic testing is crucial not only to guide treatments but screening for co-morbidities is essential for the management of patients with MODY.

The pathogenic variants and VUS uncovered in our study are consistent with that in present literature. Similar to other studies, we also found that HNF1A and GCK were the most common genes identified in our patient population with MODY^1,27^. However, a sizeable number of patients were diagnosed with variants in HNF1B. Most of these patients were diagnosed with MODY due to the presence of renal cysts noted *in utero* or in early infancy prior to the onset of diabetes. Other anomalies reported in the literature in patients with HNF1B pathogenic variants include urogenital tract deformities, hypomagnesemia, gout, and hyperparathyroidism^28^. Deletions involving a segment of chromosome 17q12 are notably prevalent among patients within the HNF1B patient cohort. This multisystem phenotype due to variants or absence of HNF1B likely contributes to the larger percentage of patients with HNF1B MODY in our patient population as well as the rapid diagnosis of MODY before onset of diabetes. This offers an opportunity for close monitoring for diabetes onset, with the potential of detecting symptoms of diabetes before patients experience sequelae of severe hyperglycemia. Finally, several rare variants were also uncovered in genes including APPL, BLK, PDX1, RFX6, CNOT1, PAX4, and NEUROD1; additional co-morbidities in these patients included pancreatic agenesis with exocrine insufficiency. Thus, timely genetic diagnosis is not only important for the appropriate treatment of diabetes but also would allow for effective management of complications in this patient population^11^.

Many patients with MODY in our study were not on medications at initial diagnosis and/or at follow-up between 1-9 months. This may reflect the fact that many of the patients in our study had GCK variants, as is the typical distribution seen in literature^1,3,8,29^. Almost all patients with variants in INS were treated with insulin, as pathogenic variants in the insulin gene lead to defects in stability of the insulin protein and its folding ultimately leading to insulin dependence^25^. Most patients with HNF1B variants were also not on medications as they were diagnosed with MODY based on genetic testing and had not yet met criteria for diabetes. Sulfonylureas were used mostly in the cohort of patients with HNF1A variants, and it has previously been reported that patients with HNF1A respond well to sulfonylureas^30,31^.

Treatment with sulfonylurea or insulin has been associated with weight gain^4,32,33^. Even though MODY has typically been reported to occur in leaner patients, the current rise in obesity could certainly be affecting patients with MODY. In a 1990 review of MODY, it was noted that patients with MODY had a higher frequency of obesity than the general population at that point in time^34^. However, obesity rates in the general population have rapidly risen since that time frame. In our patient cohort we noted an average normal BMI, but there were patients with BMI as high as 35.4 kg/m^2^. GLP-1 receptor agonists provide various metabolic advantages and could potentially be an excellent treatment option in patients with MODY, addressing both hyperglycemia and weight. In patients treated with GLP-1 receptor agonist therapy, we noted a reduction in HbA1c and the greatest reduction in BMI (−4.79%). These data indicate that future exploration of GLP-1 receptor agonist therapy in a patient population with MODY is warranted. While it is not fully understood how GLP-1 receptor agonist therapy work, pathogenic variants leading to MODY occur in transcription factors, which could affect the development of enteroendocrine cells on which GLP-1 and incretins exert effects. This could potentially enhance insulin secretion. Another study has proposed that GLP-1 receptor agonists could stimulate secretion of insulin by bypassing the decreased ATP production caused by HNF1A variants in beta cells^13–15^.

Limitations of our study include limitations in power for some of the rarer genetic variants (including CNOT1, APPL1, BLK) and genetic heterogeneity within treatment groups precluding our ability to make definitive conclusions regarding best treatment. Second, as this study was a retrospective chart review, follow-up times for comparison of BMI and HbA1c on various medications were not uniform due to patients’ availability for appointments. The racial demographics of our patient population reflected a majority of non-Hispanic white patients. While MODY has been reported to be more prevalent within this race/ethnic group, this could bias the decision to obtain screening for MODY, leading to disparities within the screening process. Finally, lack of information on diet, exercise, medication adherence, and insurance coverage of medications prohibits the ability to account for these when examining changes in BMI and HbA1c. However, strengths of this study include phenotypic characterization of patients with genetically confirmed MODY and cataloging VUS in our population with MODY. Our study provides support for identifying patients with MODY and future exploration of GLP-1 receptor agonist therapy in this patient population.

## Data Availability

All data produced in the present study are available upon reasonable request to the author

## Funding disclosure

None to disclose

## Disclosure statement

The authors do not have any duality of interest to disclose

## Acknowledgements

We would like to acknowledge the Division of Endocrinology at Cincinnati Children’s Hospital Medical Center for providing medical care, identifying patients, and aiding in the genetic testing of patients with MODY.

## Data availability

Data supporting the reported results can be obtained upon request of the authors.

## Funding

This research received no specific grant from any funding agency in the public, commercial or not-for-profit sectors.

## Authors’ Relationships and Activities

The authors have no duality of interest to disclose.

## Contribution Statement

L.D. and M.K. researched data, contributed to discussion, and wrote, reviewed, and edited the manuscript. A.S. contributed to discussion, reviewed, and edited the manuscript. All authors approved the final version of the manuscript. L.D. and M.K. are the guarantors of this work and, as such, had full access to all the data in the study and take responsibility for the integrity of the data and the accuracy of the data analysis.

---

**Supplement Figure 1.**
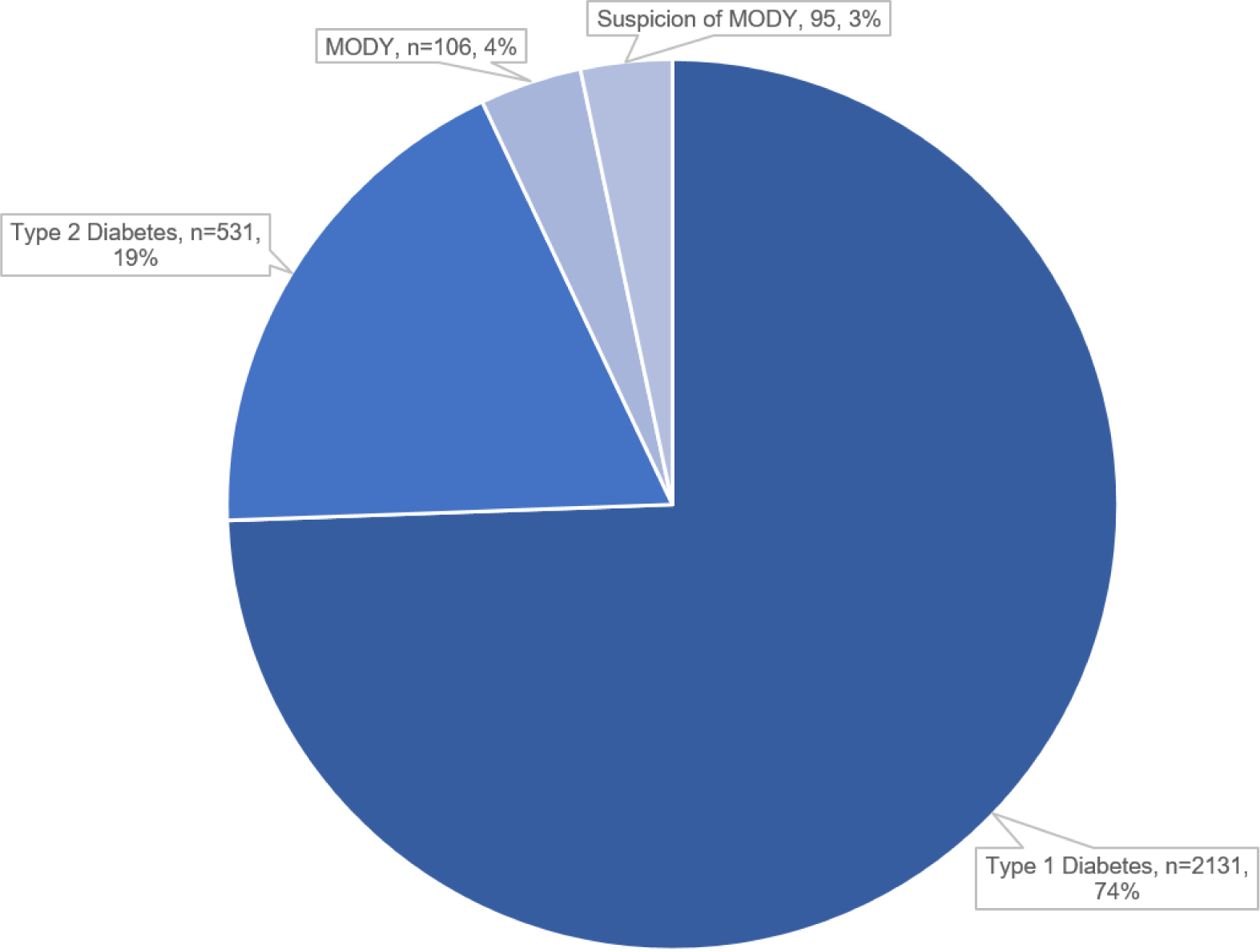
Percentage of MODY patients compared to total population with diabetes.

**Supplement Table 1.** List of all pathogenic variants and variants of unknown significance by gene.

| Genetic Variant | Zygosity | Inheritance | Significance |
| --- | --- | --- | --- |
| **ABCC8** |  |  |  |
| c.4174T>G, p.Phe1392Val | Heterozygous | AD/AR | Uncertain |
| c.291-3C>T (intronic) | Heterozygous |  | Uncertain |
| c.208G>A, p.Gly70Arg | Heterozygous | AD/AR | Pathogenic |
| c.4178 G>A, p.Arg1393His | Heterozygous | AD/AR | Likely Pathogenic |
| c.4563G>T, p.Lys1521Asn | Heterozygous | AD/AR | Uncertain |
| c.1063G>A, p.Ala355Thr | Heterozygous | AD/AR | Uncertain |
| **APPL1** |  |  |  |
| c.876 G>A, p.(L292=) | Heterozygous |  | Uncertain |
| c.682C>T, p.Pro228Ser | Heterozygous | AD | Uncertain |
| c.922A>G, p.Ile308Val | Heterozygous | AD | Uncertain |
| c.911 C>T, p.Thr304Ile | Heterozygous | AD | Uncertain |
| **BLK** |  |  |  |
| c.682C>T, p.Pro228Ser | Heterozygous | AD | Uncertain |
| c.922A>G, p.Ile308Val | Heterozygous | AD | Uncertain |
| c.911C>T, p.Thr304Ile | Heterozygous | AD | Uncertain |
| **CNOT1** |  |  |  |
| c.1459G>A, p.Ala487Thr | Heterozygous | AD | Uncertain |
| **GCK** |  |  |  |
| c.835G>C, p.Glu279Gln | Heterozygous | AD/AR | Uncertain |
| c.1351del, p.Leu451Trpfs*163 | Heterozygous | AD/AR | Pathogenic |
| c.572G>A, p.Arg191Gln | Heterozygous | AD | Likely Pathogenic |
| c.463A>G, p.Arg155Gly | Heterozygous |  | Likely Pathogenic |
| c.988T>G p.Phe330Val | Heterozygous |  | Uncertain |
| c.632T>A, p.Ile211Asn | Heterozygous | AD | Uncertain |
| c.1268T>C, p.Phe423Ser | Heterozygous |  | Uncertain |
| c.463A>G, p.Arg155Gly | Heterozygous |  | Likely Pathogenic |
| c.616A>C, p.Thr206Pro | Heterozygous | AD | Pathogenic |
| c.608T>C, p.Val203Ala | Heterozygous |  | Pathogenic |
| c.401T>G, p.Leu134Arg | Heterozygous |  | Likely Pathogenic |
| c.106C>T, p.R36W, chr7: g.44193002G>A | Heterozygous |  | Pathogenic |
| 7p13(44183089_44186518)x1 deletion (~3.43kb) | Heterozygous |  | Pathogenic |
| c.556C>T exon5, Arg186X | Heterozygous |  | Pathogenic |
| c.748C>T, p.Arg250Cys | Heterozygous | AD/AR | Likely Pathogenic |
| p.A11T c.31G.A |  |  | Polymorphism |
| c.370G>A, p.Asp124Asn | Heterozygous |  | Likely Pathogenic |
| c.883G>T, p.Gly295Cys | Heterozygous | AD | Possibly Pathogenic |
| c.1253+8C>T | Heterozygous |  | Benign |
| c.883G>T exon 8, p.Gly295Cys | Heterozygous |  | Likely Pathogenic |
| c.926T>C, p.Leu309Pro | Heterozygous |  | Pathogenic |
| c.1019G>A, p.Ser340Asn | Heterozygous |  | Uncertain |
| c.1253+8C>T | Heterozygous |  | Not Associated |
| c.1351del, p.(L451Wfs*163) | Heterozygous | AD/AR | Likely Pathogenic |
| c.871 A>T | Heterozygous | AD | Pathogenic |
| p.Lys291X | Heterozygous | AR | Pathogenic |
| c.1244T>C, p.Leu415Pro | Heterozygous | AD/AR | Uncertain |
| c.1003del, p.Val335Cysfs*18 | Heterozygous | AD/AR | Pathogenic |
| **HNF1A** |  |  |  |
| c.676_678del, p.Lys226del | Heterozygous | AD | Likely Pathogenic |
| c.599G>A, p.Arg200Gln | Heterozygous |  | Pathogenic |
| c.872dupC, P291fsinsC |  |  |  |
| c.511C>T, p.Arg171* | Heterozygous |  | Pathogenic |
| c.-160_-154dupTGGGGGT |  |  | Uncertain |
| c.452G>A, p.Gly151Asp | Heterozygous |  | Likely Pathogenic |
| c.92G>A, p.Gly31Asp | Heterozygous |  | Uncertain |
| c.787C>T, p.Arg263Cys | Heterozygous | AD | Pathogenic |
| c.1720A>G, p.Ser574Gly | homozygous |  | Not Associated |
| c.51C>G, p.Leu17Leu | Heterozygous |  | Not Associated |
| c.79A>C, p.Ile27Leu | Heterozygous |  | Not Associated |
| c.511C>T, p.Arg171* |  |  | Pathogenic |
| c.-3_25: 28 bp duplication codon 9 | Heterozygous | AD | Uncertain |
| c.787C>T, p.Arg263Cys |  |  | Likely Pathogenic |
| c.511 C>T, p.Arg171X | Heterozygous |  | Pathogenic |
| c.1084C>A, p.Leu362Met | Heterozygous |  | Possibly Pathogenic |
| c.293C>T, p.Ala98Val | Heterozygous |  | Very Unlikely<br>Pathogenic |
| c.872dupC, p.Gly292Argfs*25 | Heterozygous |  | Pathogenic |
| c.629C>T, p.Ser210Phe | Heterozygous |  | Uncertain |
| c.814 C>T, p.Arg272Cys | Heterozygous |  | Uncertain |
| c.92G>A, p.Gly31Asp | Heterozygous |  | Uncertain |
| c.1340C>T, p.Pro447Leu | Heterozygous | AD/AR | Pathogenic |
| c.-160_-154dupTGGGGGT, variant in the promoter<br>region of HNF1A |  |  | Uncertain |
| c.511C>T, p.Arg171* | Heterozygous |  | Pathogenic |
| c.1720A>G, p.Ser574Gly | homozygous |  | Not Associated |
| c.864G>C, p.Gly288Gly | Heterozygous |  | Not Associated |
| c.51C>G, p.Leu17Leu | Heterozygous |  | Not Associated |
| c.872dupC, P291fsinsC |  |  |  |
| c.155_156delinsCT, p.Gly52Ala | Heterozygous | AD/AR | Uncertain |
| c.872dup, p.G292Rfs*25 | Heterozygous | AD | Pathogenic |
| **HNF1B** |  |  |  |
| 17q12(34822500_36248918)x1 |  | Not Determined | Pathogenic |
| chr17(21562813-36248859)x1 (microdeletion including HNF1B) |  |  | Pathogenic |
| ish del(17)(q12q12)(RP11-143E18-)mat |  |  | Pathogenic |
| c.892 A>G, p. Asn298Asp | Heterozygous |  | Uncertain |
| 17q12(34815551_36244358)x1 | Heterozygous |  | Pathogenic |
| c.1045+2T>A, GT Donor | Heterozygous | AD | Likely Pathogenic |
| c.884G>A, p.Arg295His | Heterozygous | AD | Likely Pathogenic |
| 17q12(34,815,551-36,307,189)x1 |  |  | Pathogenic |
| del(17)(q12), chr17:g.34842466_36104935del | Heterozygous |  | Pathogenic |
| c.883 C>T, p.Arg295Cys | Heterozygous | AD | Likely Pathogenic |
| **HNF4A** |  |  |  |
| c.200G>A, p.Arg67Gln | Heterozygous |  | Likely Pathogenic |
| c.379C>T, p.Arg127Trp |  |  |  |
| c.925 C>T, p.Arg309Cys | Heterozygous | AD | Uncertain |
| c.908G>A, p.Arg303His | Heterozygous |  | Pathogenic |
| c.116-5C>T (No associated AA change) | Heterozygous |  | Not Associated |
| c.224+17dupT | Heterozygous |  | Less Likely Associated |
| c.724G>A, p.Val242Met | Heterozygous |  | Uncertain |
| **INS** |  |  |  |
| c.163C>T, p.Arg55Cys | Heterozygous |  | Pathogenic |
| c.-160_-143delins | Heterozygous | AD/AR | Likely Pathogenic |
| c.94G>A | Heterozygous |  | Pathogenic |
| c.140G>C, p.Gly47Ala | Heterozygous | AD/AR | Uncertain |
| c.188-31G>A | Heterozygous | AD/AR | Pathogenic |
| **KCNJ11** |  |  |  |
| c.80G>A, p.Arg27His | Heterozygous | AD/AR | Uncertain |
| c.848T>A, p.Ile283Asn | Heterozygous | AD/AR | Uncertain |
| c.190 G>T, p.Val64Leu | Heterozygous | AD/AR | Uncertain |
| **NEUROD1** |  |  |  |
| c.610C>T, p.Pro204Ser | Heterozygous | AD/AR | Uncertain |
| **PAX4** |  |  |  |
| c.776C>T, p.Ala259Val | Heterozygous | AD | Uncertain |
| **PDX1** |  |  |  |
| c.226G>A, p.Asp76Asn | homozygous | Dominant | Uncertain |
| c.349C>A, p.Leu117Met | Heterozygous | AD/AR | Uncertain |
| c.188delC(p.P63fs) | homozygous |  | Pathogenic |
| c.52T>C, p.Cys18Arg | Heterozygous | AD/AR | Uncertain |
| c.726_728dupGC, p.Pro243dup | Heterozygous | AD | Likely Pathogenic |
| c.162G>A, p.Leu54Leu | Heterozygous |  | Not Likely<br>Associated |
| **RFX6** |  |  |  |
| c.1040_1052del, p.Arg347Lysfs*18 | Heterozygous | AR | Likely Pathogenic |
| c.2623C>T, p.Gln875* | Heterozygous | AR | Likely Pathogenic |
| c.2724A>C, p.Glu908Asp | Heterozygous |  | Uncertain |
| c.1291G>A, p.Gly431Ser | Heterozygous | AD/AR | Uncertain |
| **WFS1** |  |  |  |
| c.1265C>T, p.Ala422Val | Heterozygous |  | Uncertain |
| c.1192 G>A, p.Gly398Ser | Heterozygous | AD/AR | Uncertain |
| c.991T>A, p.Phe331Ile | Heterozygous | AD/AR | Uncertain |
| c.2666C>T, p.Ala889Val | Heterozygous | AD/AR | Uncertain |
| c.1024G>A, p.Ala342Thr | Heterozygous | AD/AR | Uncertain |

**Supplement Table 2.** List of all pathogenic, likely pathogenic, uncertain significance, and not associated variants based on gene.

| Gene | Number<br>of<br>variants | Not<br>Associated | Uncertain | Likely<br>Pathogenic | Pathogenic | Polymorphism | Unknown |
| --- | --- | --- | --- | --- | --- | --- | --- |
| ABCC8 | 6 | 0 | 4 | 1 | 1 | 0 | 0 |
| APPL1 | 4 | 0 | 4 | 0 | 0 | 0 | 0 |
| BLK | 3 | 0 | 3 | 0 | 0 | 0 | 0 |
| CNOT1 | 1 | 0 | 1 | 0 | 0 | 0 | 0 |
| GCK | 28 | 2 | 6 | 9 | 10 | 1 | 0 |
| HNF1A | 30 | 7 | 8 | 4 | 9 | 0 | 2 |
| HNF1B | 10 | 0 | 1 | 3 | 6 | 0 | 0 |
| HNF4A | 7 | 1 | 3 | 1 | 1 | 0 | 1 |
| INS | 5 | 0 | 1 | 1 | 3 | 0 | 0 |
| KCNJ11 | 3 | 0 | 3 | 0 | 0 | 0 | 0 |
| NEUROD1 | 1 | 0 | 1 | 0 | 0 | 0 | 0 |
| PAX4 | 1 | 0 | 1 | 0 | 0 | 0 | 0 |
| PDX-1 | 6 | 1 | 3 | 1 | 1 | 0 | 0 |
| RFX6 | 4 | 0 | 2 | 2 | 0 | 0 | 0 |
| WFS1 | 5 | 0 | 5 | 0 | 0 | 0 | 0 |
|  |  | *Includes<br>less likely<br>associated |  | *Includes<br>Possibly<br>pathogenic |  |  |  |

## Notes

### Competing Interest Statement

The authors have declared no competing interest.

### Funding Statement

This study did not receive any funding.

### Author Declarations

IRB of Cincinnati Children's Hospital Medical Center gave ethical approval for this work.

